# A comparison of paediatric hypertension clinical practice guidelines and their ability to predict adult hypertension in an African birth cohort

**DOI:** 10.1101/2022.03.22.22272744

**Authors:** Ashleigh Craig, Lisa J Ware, Witness Mapanga, Shane A Norris

## Abstract

It remains unclear which paediatric hypertension clinical practice guideline (CPG) should be applied in an African population. We therefore aimed to compare three commonly used CPG (2017 AAP, 2016 ESH and 2004 Fourth Report) developed in high-income countries for use in South African children at four paediatric ages (children: 5yrs and 8yrs; adolescents: 13yrs and 17yrs) to determine which best predicts elevated blood pressure (EBP) in young adulthood (22yrs and 28yrs). Moreover, the sensitivity, specificity, positive predictive value (PPV) and negative predictive value (NPV) for each specific paediatric CPG was calculated. The 2017 AAP definition identified more children and adolescents with hypertension when compared to the 2004 Fourth Report and 2016 ESH guidelines. In computed hazards ratios, from ages 8yrs to 17yrs, all three paediatric CPG significantly predicted the risk of EBP in young adulthood (p≤0.008). However, sensitivity to predict EBP at age 22yrs for all CPG was generally low (17.0% - 33.0%) with higher specificity (87.4% - 93.1%). Sensitivity increased at age 28yrs (51.4.0% - 70.1%), while specificity decreased (52.8% - 65.1%). Both PPV and NPV at both adult age points varied widely (17.9% - 79.9% and 29.3% - 92.5% respectively). The performance of these paediatric CPG in terms of AUC were not optimal at both adult age points, however, the AAP definition at 17yrs met an acceptable level of performance (AUC= 0.71). Our results highlight the need for more research to examine if an African-specific CPG would better identify high-risk children to minimise their trajectory towards adult hypertension.

## Introduction

Hypertension (HTN) is a renowned modifiable risk factor for the development of cardiovascular disease (CVD) [1], that is established early in life [2] and affects over 1 billion individuals globally [3]. Until recently, HTN affected mainly affluent regions of the world, however, low and middle-income countries (LMICs) now account for two-thirds of the global prevalence, which may be explained by inadequate identification, prevention and treatment strategies [4].

Epidemiologically designed longitudinal studies have shown childhood blood pressure (BP) tracks into adulthood [5-7]. It is therefore necessary to identify those young individuals who are at high-risk in order to implement the necessary intervention strategies for the early prevention of CVD. In recognising this, the Fourth Report on the Diagnosis, Evaluation, and Treatment of High Blood Pressure in Children and Adolescents (NHBPEP) (2004 Fourth report) [8], the American Academy of Pediatrics (2017 AAP) [9] and the European Society of Hypertension (2016 ESH) [10] among others, have provided age, sex and height specific BP guidelines to allow for the identification of high-risk children.

With the increasing interest surrounding the effect of childhood HTN and CV risk in later life, evidence has revealed that HTN in children is not as uncommon as previously believed, and in most cases represents CV risk in young adulthood [11]. Adolescents with previous high childhood BP are also more likely to present with persistent HTN in adulthood [12]. Furthermore, hypertensive youth are also susceptible to accelerated vascular aging [13] resulting in premature structural and functional alteration within the arterial wall enhancing early onset CVD [14]. Thus, the identification and treatment of early onset HTN is paramount.

Several population-based studies in several countries (including China, India, Iran, Korea, Poland, Tunisia and United States (US)), have prospectively examined the usefulness of these high-income counties (HICs) paediatric clinical practice guidelines (CPG) in identifying high-risk individuals [15, 16]. Certainly, adult HTN CPG are frequently applied in LMIC settings. For example, systolic BP (SBP) above 140 mmHg (as commonly applied in North America and the European region) has been shown to associate with 6-year mortality in adults from a LMIC [17], thus illustrating that a HICs BP guideline can perform in LMIC adult populations. Whether this can also be replicated in children and more specifically in African children, is unclear and warrants further exploration. We therefore aimed to compare three commonly used childhood CPG at four paediatric ages (children 5yrs and 8yrs; adolescents 13yrs and 17yrs) to determine which best predicts elevated blood pressure (EBP) in adulthood (22yrs and 28yrs) in a South African cohort.

## Methodology

The longitudinal Birth-to-Twenty Plus (Bt20) Cohort enrolled 3273 babies born between April and June 1990 in Soweto-Johannesburg (South Africa) and who remained residents in the area for 6 months. Socioeconomic, personal and familial information pertaining to physical and psychological health and well-being have now been recorded 23 times since birth. The study population and protocol are described elsewhere [18]. Briefly, the study was designed to assess young childhood and adolescent growth, development and overall health, following the demographic transition in South Africa.

For this study, BP data that was collected in childhood (5yrs; *n*= 1235 and 8yrs; *n*= 1321), adolescence (13yrs; *n*= 1619 and 17yrs; *n*= 1853) and adulthood (22yrs; *n*= 1541 and 28yrs; *n*=917) were utilised. The study was conducted in line with the ethical principles of the Declaration of Helsinki [19] and obtained approval from the Human Research Ethics Committee (Medical) of the University of the Witwatersrand (South Africa) (Ref: M190263). All participants were fully informed about the objectives of the study and written informed consent/assent was obtained from each adult and child participant respectively.

### Measurements

Height in cm was measured with a stadiometer (Holtain, United Kingdom) according to specific guidelines and collected using standard protocols [20]. Prior to the measurement of BP, participants were required to remain in a seated position with uncrossed legs for 5 minutes. Three BP measurements were obtained at age 5yrs using the automated Dinamap Signs monitor 1846SX (Critikon, USA) and at age 8yrs and above using an OMRON M6 oscillometric BP monitor (OMRON HealthCare Co., LTD. Kyoto, Japan). Both devices have been validated for use in children [21]. Using an appropriately sized BP cuff, brachial BP was measured on the left upper arm at 2-minute intervals. The first BP measure was discarded while, the mean of the second and third measures were used for. Systolic BP (SBP) and diastolic BP (DBP) were captured from each measurement. Mean arterial pressure (MAP) was subsequently calculated using the formula: [(2 x DBP) + SBP] / 3 [22].

Blood pressure status of the children and adolescent participants (normotensive, pre-HTN and HTN) was classified using specific age, sex and body height reference standards according to the three paediatric CPG (2004 Fourth report [8], 2017 AAP [9] and 2016 ESH [10]; **Table 1**). Adult BP status was classified using the South African hypertension practice guideline 2014 (normal: <120mmHg/<80mmHg; pre-HTN:130-139mmHg/80-85mmHg and HTN: >140mmHg/>90mmHg) [23]. For adverse BP as an outcome in adulthood, pre-HTN and HTN were grouped as EBP.

**Table 1.** Childhood, and adolescent clinical practice guidelines (5-17yrs).

| Classification guideline | Normal | Pre-hypertension | Hypertension<br>Stage 1 | Hypertension<br>Stage 2 |
| --- | --- | --- | --- | --- |
| The Fourth Report on the Diagnosis, Evaluation, and Treatment of High Blood Pressure in Children and Adolescents [8]. | 1–13yrs: Average brachial SBP and/or DBP <90<sup>th</sup> percentile for age, sex and height. | 1–13yrs: Average brachial SBP and/or DBP ≥90<sup>th</sup> percentile but <95<sup>th</sup> percentile. | 1–17yrs: Average brachial SBP and/or DBP ≥95<sup>th</sup> percentile to <95<sup>th</sup> percentile + 12mmHg for age, sex and height (on 3 occasions). | 1–17rs: Average brachial SBP and/or DBP ≥95<sup>th</sup> percentile +12mmHg for age, sex and height (on 3 occasions). |
| American Academy of Pediatrics “Clinical Practice Guideline for Screening and Management of High Blood pressure in Children and Adolescents.” [9]. | <13 yrs: Average brachial SBP and/or DBP <50<sup>th</sup> percentile for age, sex and height. Separate chart for gender. Actual height in cm or inches is provide on chart.<br>≥13yrs: <120/<80mmHg | <13 yrs: Average brachial SBP and/or DBP <90<sup>th</sup> percentile for age, sex and height. Separate chart for gender. Actual height in cm or inches is provide on chart.<br>≥13yrs: 120/<80mmHg to 129-80 mmHg. | <13 yrs: Average brachial SBP and/or DBP ≥95<sup>th</sup> percentile for age, sex and height. Separate chart for gender. Actual height in cm or inches is provide on chart.<br>≥13yrs: 130/80mmHg to 139/89 mmHg. | <13 yrs: Average brachial SBP and/or DBP ≥95<sup>th</sup> percentile +12mmHg for age, sex and height. Separate chart for gender. Actual height in cm or inches is provide on chart.<br>≥13yrs: ≥140/90mmHg |
| 2016 European Society of Hypertension guidelines for the management of high blood pressure in children and adolescents [10]. | <16 yrs: Brachial SBP and/or DBP <90<sup>th</sup> percentile.<br>≥16 yrs: Brachial SBP and/or DBP 130/85mmHg. | <16 yrs: Brachial SBP and/or DBP ≥90<sup>th</sup> to 95<sup>th</sup> percentile.<br>≥16 yrs: Brachial BP 130/85mmHg to 139/89 mmHg. | 0-15yrs: Brachial SBP and/or DBP 95<sup>th</sup> to 99<sup>th</sup> percentile + 5mmHg.<br>≥16 yrs: Brachial BP 140/90mmHg to 159/99mmHg. | <16 yrs: Brachial SBP and/or DBP >99<sup>th</sup> percentile + 5mmHg.<br>≥16 yrs: Brachial BP 160/100mmHg to 179/109mmHg. |

## Statistical analyses

All statistical analyses were performed using IBM^®^ SPSS^®^ version 27 (IBM Corporation, Armonk, New York). Categorical variables were presented as frequency and percentage. Proportions were determined with cross-tabs with significant differences indicated by Chi-square tests. Hazard ratios were computed at each age interval to predict which paediatric CPG best predicts EBP in adulthood. Furthermore, the sensitivity, specificity, positive predictive value (PPV) and negative predictive value (NPV) for each specific paediatric CPG was calculated at specific age intervals. Additionally, as performance measures were assessed across various age points, the area under the receiver operating characteristic curve (AUC) [ranging from 0.5 (fail) to 1.0 (excellent)] was measured to reflect the accuracy of the specific paediatric CPG to predict those adults with and without EBP as an outcome.

## Results

Overall, each age interval presented with an equal sex distribution (males: 5yrs: *n*=611 (49.5%); 8yrs: *n*=651 (49.3%); 13yrs: *n*=772 (47.7%); 17yrs: *n*=888 (47.9%); 22yrs: *n*=755 (49.0%) and 28yrs: *n*=442 (48.2%)). The prevalence of normotension, pre-HTN and HTN at various age intervals according to each guideline was derived (**Table 2a** and **2b**). At age 5yrs the 2017 AAP classification gave a >1.07-fold higher number of hypertensive children than the Fourth 2004 report and ESH 2016 guidelines. At age 8yrs the 2017 AAP classification still gave a >1.23-fold higher number of hypertensive children than the two other CPG. Likewise in adolescence, this difference was greater with a >3.33-fold difference using the 2017 AAP guideline compared to the 2004 Fourth report. In adulthood, pre-HTN and HTN prevalence were 9.7% and 5.4% at age 22yrs and 36.5% and 12.6% at age 28yrs respectively.

**Table 2a.** Blood pressure status of children and adolescence stratified according to mean age intervals based on specific paediatric clinical practice guidelines (*n* (%)).

|  | Normal | Pre-HTN | HTN |
| --- | --- | --- | --- |
| **Guideline 1 – Fourth Report (2004)** |  |  |  |
| Age 5yrs (*n*= 1235) | 517 (41.9%) | 361 (29.2%) | 357 (28.9%) |
| Age 8yrs (*n*= 1321) | 598 (45.3%) | 473 (35.8%) | 250 (18.9%) |
| Age 13yrs (*n*= 1619) | 1304 (80.5%) | 265 (16.4%) | 49 (3.1%) |
| Age 17yrs (*n*= 1853) | 1279 (69.0%) | 475 (24.7%) | 99 (5.3%) |
| **Guideline 2 – AAP (2017)** |  |  |  |
| Age 5yrs (*n*= 1235) | 359 (29.0%) | 282 (22.8%) | 594 (48.0%) |
| Age 8yrs (*n*= 1321) | 425 (32.2%) | 289 (21.8%) | 607 (46.0%) |
| Age 13yrs (*n*= 1619) | 1409 (87.0%) | 50 (3.0%) | 160 (10.0%) |
| Age 17yrs (*n*= 1853) | 1070 (57.7%) | 421 (22.7%) | 362 (19.5%) |
| **Guideline 3 – ESH (2016)** |  |  |  |
| Age 5yrs (*n*= 1235) | 530 (42.9%) | 153 (12.4%) | 552 (44.7%) |
| Age 8yrs (*n*= 1321) | 560 (42.4%) | 267 (20.2%) | 494 (37.4%) |
| Age 13yrs (*n*= 1619) | 1293 (79.9%) | 153 (9.5%) | 172 (10.6%) |
| Age 17yrs (*n*= 1853) | 1558 (84.1%) | 213 (11.5%) | 82 (4.4%) |

Abbreviations: *n* – number of participants; pre-HTN – pre-hypertension; HTN – hypertension; Fourth report - The Fourth Report on the Diagnosis, Evaluation, and Treatment of High Blood Pressure in Children and Adolescents; AAP – American Academy of Pediatrics and; ESH – European Society of Hypertension.

**Table 2b.** Blood pressure status in adulthood of the study population based on the South African hypertension practice guideline 2014 (*n* (%)).

|  | Normal | Pre-HTN | HTN |
| --- | --- | --- | --- |
| Age 22yrs (*n*= 1540) | 1308 (84.9%) | 150 (9.7%) | 83 (5.4%) |
| Age 28yrs (*n*= 917) | 467 (50.9%) | 335 (36.5%) | 115 (12.6%) |

Abbreviations: *n* – number of participants; pre-HTN – pre-hypertension; HTN – hypertension.

In determining which paediatric CPG at each age best predicted EBP in adulthood (age 22yrs and 28yrs) (**Table 3a** and **3b**), we found that at all paediatric ages, except for age 5yrs (*p*≥0.051), all three of the paediatric CPG significantly predicted EBP in adulthood at both age 22yrs (*p*≤0.008) and 28yrs (*p*≤0.001). Overall, with EBP as an outcome at age 22yrs, the hazard ratio increased with age in each specific CPG, showing a larger hazards ratio with more significance in older participants (age 17yrs). At different paediatric ages, different CPG performed better in predicting adult EBP. At age 8 years, the 2017 AAP guideline showed the best prediction of adult (28yrs) EBP with adverse BP at age 8yrs (β= 1.40; [95% CI 1.22; 1.61]; *p*<0.001). At age 13yrs, the 2016 ESH guideline showed the best prediction of adult (28yrs) EBP (β= 1.27; [95% CI 1.16; 1.39]; *p*<0.001), while the 2004 Fourth report showed the best prediction of adult (28yrs) EBP (β= 1.29; [95% CI 1.18; 1.42]; *p*<0.001) at age 17yrs respectively.

**Table 3a.** Multivariate-adjusted hazards ratio to predict which paediatric clinical practice guideline at each age interval best predicts elevated blood pressure as an outcome at age 22yrs.

| | $\beta$ | 95% (CI) | *p* value |
| --- | --- | --- | --- |
| **Guideline 1 – Fourth Report (2004)** |  |  |  |
| Age 5yrs (*n*= 832) | 1.13 | (0.95; 1.36) | 0.18 |
| Age 8yrs (*n*= 879) | 1.36 | (1.33; 1.64) | **0.001** |
| Age 13yrs (*n*= 1125) | 1.57 | (1.30; 1.83) | **<0.001** |
| Age 17yrs (*n*= 1422) | 1.62 | (1.43; 1.85) | **<0.001** |
| **Guideline 2 – AAP (2017)** |  |  |  |
| Age 5yrs (*n*= 832) | 1.32 | (1.06; 1.64) | 0.10 |
| Age 8yrs (*n*= 879) | 1.39 | (1.07; 1.59) | **0.008** |
| Age 13yrs (*n*= 1125) | 1.45 | (1.15; 1.61) | **<0.001** |
| Age 17yrs (*n*= 1422) | 1.76 | (1.51; 2.06) | **<0.001** |
| **Guideline 3 – ESH (2016)** |  |  |  |
| Age 5yrs (*n*= 832) | 1.21 | (1.00; 1.45) | 0.051 |
| Age 8yrs (*n*= 879) | 1.24 | (1.03; 1.49) | **0.021** |
| Age 13yrs (*n*= 1125) | 1.38 | (1.22; 1.56) | **<0.001** |
| Age 17yrs (*n*= 1422) | 1.53 | (1.32; 1.79) | **<0.001** |

Variables included in the models were: sex and childhood BMI z-score. Abbreviations: *n* – number of participants; Fourth report - The Fourth Report on the Diagnosis, Evaluation, and Treatment of High Blood Pressure in Children and Adolescents; AAP – American Academy of Pediatrics and; ESH – European Society of Hypertension. Bold values denote statistical significance (*p*<0.05).

**Table 3b.** Multivariate-adjusted hazards ratio to predict which paediatric clinical practice guideline at each age interval best predicts elevated blood pressure as an outcome at age 28yrs.

| | $\beta$ | 95% (CI) | *p* value |
| --- | --- | --- | --- |
| **Guideline 1 – Fourth Report (2004)** |  |  |  |
| Age 5yrs (*n*= 496) | 0.98 | (0.86; 1.11) | 0.76 |
| Age 8yrs (*n*= 525) | 1.36 | (1.19; 1.55) | **<0.001** |
| Age 13yrs (*n*= 764) | 1.25 | (1.14; 1.37) | **<0.001** |
| Age 17yrs (*n*= 863) | 1.29 | (1.18; 1.42) | **<0.001** |
| **Guideline 2 – AAP (2017)** |  |  |  |
| Age 5yrs (*n*= 496) | 1.06 | (0.93; 1.21) | 0.35 |
| Age 8yrs (*n*= 525) | 1.40 | (1.22; 1.61) | **<0.001** |
| Age 13yrs (*n*= 764) | 1.21 | (1.10; 1.32) | **<0.001** |
| Age 17yrs (*n*= 863) | 1.28 | (1.16; 1.42) | **<0.001** |
| **Guideline 3 – ESH (2016)** |  |  |  |
| Age 5yrs (*n*= 496) | 1.01 | 0.89; 1.14) | 0.94 |
| Age 8yrs (*n*= 525) | 1.25 | (1.10; 1.42) | **0.001** |
| Age 13yrs (*n*= 764) | 1.27 | (1.16; 1.39) | **<0.001** |
| Age 17yrs (*n*= 863) | 1.26 | (1.15; 1.37) | **<0.001** |

Variables included in the models were: sex and childhood BMI z-score. Abbreviations: *n* – number of participants; Fourth report - The Fourth Report on the Diagnosis, Evaluation, and Treatment of High Blood Pressure in Children and Adolescents; AAP – American Academy of Pediatrics and; ESH – European Society of Hypertension. Bold values denote statistical significance ( $p < 0.05$ ).

A comparison of the performance of the three specific CPG to identify EBP in adulthood (age 22yrs and 28yrs) is presented in **Tables 4a** and **4b**. At age 22yrs, across all CPG, the thresholds manifested a low sensitivity (17.0% - 33.0%), a high specificity (87.4% - 93.1%), a moderately low to high PPV (23.5% - 79.9%) and corresponding NPV (29.3% - 88.9%). At age 28yrs, the thresholds to identify EBP in adulthood indicated that sensitivity increased to a moderate to high range (51.4.0% - 70.1%), while specificity decreased to a moderate range (52.8% - 65.1%). The PPV (17.9% - 77.7%) and NPV (33.6% - 92.5%) at age 28yrs still remained wide spread from moderately low to high. Furthermore, at both adult outcomes (age 22yrs and 28yrs), performance of these CPG in terms of AUC suggested that only the 2017 AAP definition at age 17yrs (AUC= 0.71) met an acceptable level of performance to identify EBP in adulthood (22yrs), while all other CPG did not perform adequately in identifying EBP in adults (AUC≤0.68).

**Table 4a.** Sensitivity, specificity, predicative value and accuracy of paediatric clinical practice guideline at each age interval to predict elevated blood pressure as an outcome at age 22yrs.

|  | Guideline | Sensitivity<br>(95% CI) | Specificity<br>(95% CI) | Predictive value (95% CI) |  | AUC<br>(95% CI) | Absolute<br>adults detected<br>*n* (%) | Absolute<br>adults missed<br>*n* (%) |
| --- | --- | --- | --- | --- | --- | --- | --- | --- |
|  |  |  |  | Positive | Negative |  |  |  |
| EBP & HTN<br>outcome<br>(Age 5yrs) | Fourth (2004) | 17.7 (14.5; 21.4) | 87.8 (83.7; 91.0) | 68.2 (59.4; 76.0) | 41.8 (38.2; 45.6) | 0.53 (0.46; 0.60) | 109 (13.1) | 20 (2.4) |
|  | AAP (2017) | 18.0 (15.1; 21.3) | 91.2 (86.5; 94.4) | 84.5 (76.8; 90.0) | 29.3(26.0; 32.8) | 0.55 (0.48; 0.62) | 88 (10.6) | 41 (4.9) |
|  | ESH (2016) | 18.3 (15.0; 22.0) | 88.5 (84.5; 91.6) | 69.8 (61.0; 77.4) | 42.7 (39.0; 46.4) | 0.55 (0.48; 0.62) | 90 (10.8) | 39 (4.7) |
| EBP & HTN<br>outcome<br>(Age 8yrs) | Fourth (2004) | 20.4 (16.9; 24.3) | 89.8 (86.3; 92.5) | 71.2 (62.8; 78.4) | 47.7 (44.1; 51.4) | 0.59 (0.52; 0.66) | 99 (11.3) | 40 (4.6) |
|  | AAP (2017) | 18.3 (15.4; 21.7) | 89.8 (85.4; 93.0) | 79.9 (72.0; 86.0) | 33.2 (29.9; 36.8) | 0.59 (0.52; 0.65) | 111 (12.6) | 28 (3.2) |
|  | ESH (2016) | 19.3 (16.0; 23.0) | 89.1 (85.3; 92.0) | 71.2 (62.8; 78.0) | 44.1 (40.5; 47.7) | 0.56 (0.49; 0.63) | 100 (11.4) | 41 (4.7) |
| EBP & HTN<br>outcome<br>(Age 13yrs) | Fourth (2004) | 26.0 (20.7; 32.1) | 87.4 (85.1; 89.4) | 33.7 (27.1; 41.0) | 82.8 (80.3; 85.0) | 0.58 (0.51; 0.66) | 63 (5.6) | 124 (11.0) |
|  | AAP (2017) | 27.7 (21.0; 35.4) | 86.6 (84.4; 88.5) | 23.5 (17.9; 30.4) | 88.9 (86.8; 90.7) | 0.55 (0.47; 0.62) | 44 (3.9) | 143 (12.7) |
|  | ESH (2016) | 27.6 (22.2; 33.6) | 87.9 (85.7; 89.9) | 37.4 (30.6; 44.8) | 82.3 (79.8; 84.5) | 0.61 (0.54; 0.69) | 70 (6.2) | 117 (10.4) |
| EBP & HTN<br>outcome<br>(Age 17yrs) | Fourth (2004) | 28.8 (24.6; 33.3) | 91.1 (89.1; 92.8) | 58.7 (51.7; 65.3) | 74.4 (71.9; 78.6) | 0.68 (0.61; 0.75) | 125 (8.8) | 88 (6.2) |
|  | AAP (2017) | 26.0 (22.5; 29.7) | 93.1 (91.0; 94.7) | 73.2 (66.7; 78.9) | 63.2 (60.4; 65.9) | 0.71 (0.65; 0.78) | 156 (11.0) | 57 (4.0) |
|  | ESH (2016) | 33.0 (27.0; 40.0) | 88.4 (86.4; 90.1) | 34.7 (28.4; 41.6) | 87.6 (85.6; 79.6) | 0.59 (0.52; 0.67) | 74 (6.5) | 139 (9.8) |

Abbreviations: *n* – number of participants; AUC – area under the curve Fourth report - The Fourth Report on the Diagnosis, Evaluation, and Treatment of High Blood Pressure in Children and Adolescents; AAP – American Academy of Pediatrics and; ESH – European Society of Hypertension.

**Table 4b.** Sensitivity, specificity, predicative value and accuracy of paediatric clinical practice guideline at each age interval to predict elevated blood pressure as an outcome at age 28yrs.

|  | Guideline | Sensitivity<br>(95% CI) | Specificity<br>(95% CI) | Predictive valve (95% CI) |  | AUC<br>(95% CI) | Absolute<br>adults detected<br>*n* (%) | Absolute<br>adults missed<br>*n* (%) |
| --- | --- | --- | --- | --- | --- | --- | --- | --- |
|  |  |  |  | Positive | Negative |  |  |  |
| EBP & HTN<br>outcome<br>(Age 5yrs) | Fourth (2004) | 51.4 (45.4; 57.4) | 52.8 (45.9; 59.6) | 58.9 (52.2; 65.1) | 45.2 (38.9; 51.6) | 0.52 (0.45; 0.59) | 145 (29.2) | 101 (20.4) |
|  | AAP (2017) | 52.2 (46.8; 57.5) | 56.4 (48.0; 64.4) | 73.6 (67.5; 78.9) | 33.6 (27.8; 39.9) | 0.54 (0.47; 0.60) | 181 (36.5) | 65 (13.1) |
|  | ESH (2016) | 53.9 (47.0; 60.6) | 52.3 (46.3; 58.3) | 59.3 (52.9; 65.5) | 46.8 (40.5; 53.1) | 0.53 (0.47; 0.60) | 146 (29.4) | 100 (20.2) |
| EBP & HTN<br>outcome<br>(Age 8yrs) | Fourth (2004) | 59.1 (53.2; 64.7) | 63.0 (56.3; 69.2) | 57.7 (61.6; 73.3) | 54.0 (47.8; 60.0) | 0.60 (0.53; 0.66) | 176 (33.5) | 84 (16.0) |
|  | AAP (2017) | 56.3 (51.0; 61.4) | 65.1 (57.2; 72.2) | 77.7 (72.0; 82.5) | 40.8 (34.8; 47.0) | 0.60 (0.53; 0.66) | 202 (38.5) | 58 (11.0) |
|  | ESH (2016) | 56.7 (51.0; 62.3) | 61.0 (54.0; 67.6) | 68.1 (62.0; 73.6) | 49.1 (42.9; 55.2) | 0.57 (0.50; 0.63) | 177 (33.7) | 83 (15.8) |
| EBP & HTN<br>outcome<br>(Age 13yrs) | Fourth (2004) | 69.9 (61.6; 77.0) | 55.8 (51.8; 60.0) | 27.2 (22.8; 32.1) | 88.7 (85.0; 91.6) | 0.59 (0.52; 0.65) | 102 (13.4) | 273 (35.7) |
|  | AAP (2017) | 69.8 (59.4; 78.5) | 53.9 (50.0; 57.7) | 17.9 (14.2; 22.2) | 92.5 (89.4; 94.9) | 0.55 (0.49; 0.62) | 67 (8.8) | 308 (40.3) |
|  | ESH (2016) | 68.4 (60.4; 75.5) | 55.8 (51.7; 59.7) | 28.3 (23.8; 33.1) | 87.4 (83.6; 90.4) | 0.59 (0.52; 0.66) | 106 (13.9) | 269 (35.2) |
| EBP & HTN<br>outcome<br>(Age 17yrs) | Fourth (2004) | 66.7 (60.7; 72.2) | 59.2 (55.1; 63.1) | 43.0 (38.3; 47.9) | 79.3 (75.2; 82.9) | 0.62 (0.56; 0.69) | 182 (21.1) | 241 (27.9) |
|  | AAP (2017) | 63.0 (57.8; 67.9) | 61.5 (57.0; 65.8) | 55.1 (50.2; 59.9) | 68.9 (64.3; 73.1) | 0.61 (0.55; 0.68) | 233 (27.0) | 190 (22.0) |
|  | ESH (2016) | 70.1 (61.9; 77.3) | 55.2 (51.5; 58.9) | 23.9 (20.0; 28.3) | 90.2 (87.0; 92.8) | 0.57 (0.50; 0.64) | 101 (11.7) | 322 (37.3) |

Abbreviations: *n* – number of participants; AUC – area under the curve; Fourth report - The Fourth Report on the Diagnosis, Evaluation, and Treatment of High Blood Pressure in Children and Adolescents; AAP – American Academy of Pediatrics and; ESH – European Society of Hypertension.

## Discussion

Our aim was to compare the three most commonly applied International paediatric CPG for HTN in an African paediatric cohort and to assess the utility of these to predict EBP in adulthood. Examining the prevalence of childhood HTN, we found that there was a higher HTN prevalence using the 2017 AAP guideline than using either the 2004 Fourth Report or the 2016 ESH guidelines. Across various childhood and adolescent age points, the 2017 AAP definition indeed showed an expected higher HTN prevalence, which is consistent with the findings from numerous recent cross-sectional studies conducted worldwide [16, 24, 25]. We also found that the prevalence of pre-HTN decreased across our specific ages with a concomitant upward trend in the prevalence of HTN [15; 26]. Although contrary to our findings, one US study reported a lower HTN prevalence when applying the 2017 AAP guidelines in a large paediatric population when compared to the 2004 Fourth report, presumably due to differences in age range [27].

The most prominent reason why several studies, including ours, have observed disparities in HTN prevalence when applying the 2017 AAP definition compared with the two other paediatric definitions, could be due to the notable change seen in the revised BP charts adapted by 2017 AAP guideline, which excludes overweight and obese individuals. It is evident that higher levels of adiposity represent one of the most important risk factors for development of HTN in children [28]. This exclusion of overweight and obese children in the development of the CPG yields a lower BP cut off value (2-3 mmHg lower) when compared with the original BP charts included in the 2004 Fourth Report. Consequently, in a population sample that includes overweight and obese children and adolescents, the overall prevalence will increase.

Other likely reasons may be due to linear growth of the study population, as height is a major independent determinant of childhood BP classification [29]. Comparing the 2017 AAP and 2016 ESH guidelines, the 2017 AAP definition shows a much broader BP range in shorter individuals (those in the lower 50% height percentile), while this gap tends to fade in taller individuals (those between 90-95% height percentiles) [30]. Consequently, the application of the 2017 AAP guidelines could in theory reclassify those with a shorter stature downward into a lower BP category. When evaluating the growth of the Bt20 study population included in this study, all age points showed on average lower linear growth than that described by age, sex and mean height percentiles of the World Health Organisation (WHO) [31]. Therefore, this highlights the need to further examine the linear growth effect on childhood BP classification in an LMIC setting.

Considering childhood BP is known to track into adulthood, we further sought to determine how well paediatric CPG predict adverse BP in adults. When looking at the performance of these results, in fact, the performance of all three CPG were not optimal in childhood to predict EBP in adulthood. Additionally at age 22yrs, our performance results show using paediatric CPG were highly specific (specificity >86%) yet not sensitive (sensitivity <35%) in predicting adverse BP in adulthood, although the closer to adulthood the measurement was, the better the prediction (2017 AAP definition showed an AUC value of 0.71 at age 17yrs), which may highlight a potential window of opportunity for screening high-risk adolescents. It has been reported that juvenile BP measured in childhood and adolescence strongly predicts BP in mid-life [32] and given that the Bt20 cohort comprises of young adults (<30yrs of age), we may expect less predictability. The sensitivity of our performance did, however, moderately increase (<70%) as older adults were assessed (28yrs), but specificity at this age point decreased (<65%). Consequently, across all paediatric definitions <13% of children and adolescents who previously were classified as having childhood pre-HTN or HTN remained with adverse BP as adults at age 22yrs and; <39% at age 28yrs respectively. At older adult age points, more of the cohort exhibited pre-HTN (by +26.8%) and HTN (by +7.2%) showing a 3-fold increase in adverse BP in a span of just 6 years, which could have influenced the ability to detect and/or predict adverse BP from earlier measures.

As childhood HTN is often unrecognised due to symptoms rarely presenting among children, as in adults, and HTN-related complications usually occur several years after onset [8, 9], identifying high-risk children and adolescents allows for the opportunity to implement low-cost lifestyle changes, which may halt or even reverse the progression of HTN and reduce early onset CV alterations. However, as a result, applying the revised 2017 AAP definition has been frequently adopted worldwide, where more children and adolescents are classified as hypertensive, this simple adoption of childhood BP nomograms from a HIC such as the US may, however, may not be best suited for LMICs with diverse population characteristics. These guidelines may therefore introduce unpredicted bias in evaluating childhood BP resulting in a significant over or under-estimation of normotension, pre-HTN and/or HTN in children and adolescence. Therefore, the need to explore country or region-specific CPG has gained much focus over the past decade [33-35].

Our findings must be interpreted within the context of the strengths and limitations of this study. This study was the first to compare three commonly used paediatric CPG to assess the prevalence of childhood HTN and the risk of EBP in adulthood in an African cohort. We also acknowledge that our study was limited in the fact that BP measures were only obtained at one data collection timepoint per age interval and not on three separate occasions as per recommended guidelines. Also, out of home measurements may increase BP due to white coat effects, even though Bt20 is a birth cohort and participants are very comfortable and familiar with study staff and setting. Our study included urban African participants and may not be representative of a national sample.

In conclusion, our results showed a concerning picture of rising EBP (combined pre-HTN and HTN) prevalence from childhood into adulthood, which warrants more prevention efforts to be upscaled with appropriate methods and guidelines to support this. Also, we confirmed that the prevalence of HTN among children and adolescents was higher when the 2017 AAP guidelines were applied than those of the 2004 Fourth Report or 2016 ESH guidelines. All three specific paediatric CPG were found to significantly predict the risk of EBP in adulthood at ages 8yrs, 13yrs and 17yrs respectively. This, however, must be interpreted with caution considering the poor predictability performance of these estimates.

## Data Availability

All data produced in the present work are contained in the manuscript.

## Acknowledgments

The authors are thankful towards all participants in the Bt20 cohort study. AC, WM, LJW, SAN are supported by the NRF-DSI Centre of Excellence in Human Development at the University of the Witwatersrand, Johannesburg, South Africa. LJW is supported by the Wellcome Trust [grant 214082/Z/18/Z].

## Author contributions

All authors conceived and/or designed the work that led to the submission, revised the manuscript, approved the final version, agreed to be accountable for all aspects of the work. AC carried out the data analyses and generated tables, interpreted the data, compiled literature search, and was responsible for writing the manuscript. LJW, WM, and SAN interpreted the data and made a significant contribution in the interpretation of the results.

